# Evaluation Of A New “All In One” Sars-Cov-2 Antigen-Detecting Rapid Diagnostic Test And Self-Test: Diagnostic Performance And Usability On Child And Adult Population

**DOI:** 10.1101/2021.09.14.21263403

**Authors:** Thierry Prazuck, Anne Gravier, Daniela Pires-Roteira, Aurelie Theillay, Sandra Pallay, Mathilda Colin, Raphael Serreau, Laurent Hocqueloux, Nino Guy Cassuto

## Abstract

The control of the COVID-19 epidemics has been one of top global health priorities for the last eighteen months. To that end, more reliable and easy-to-use diagnostic tests are necessary. Young children are still not eligible to vaccination and it is important to find a way to easily test this key population regularly. With that in mind, we evaluated a new innovative easy two-step self-test named COVID-VIRO ALL IN^®^ developed by AAZ that uses a sampling nasal sponge instead of a classic nasal swab. Mirroring the previous study conducted on the first generation of COVID-VIRO^®^ antigenic self-test, we first performed a multicentre, prospective study on 124 adults and children, in a point-of-care setting. Sensitivity, specificity and overall acceptance of the COVID-VIRO ALL IN^®^ antigen self-test compared to RT-PCR on nasopharyngeal samples were evaluated at 93.02%, 100% and 97,5%, respectively. We then performed a multicentre, prospective, usability study to evaluate the ease of use of COVID-VIRO ALL IN^®^ in real life on 68 laypersons, all adults. Globally, 99% of them considered the instructions material good, 98% executed the procedure well, and all of them interpreted the results correctly. The usability was then specifically investigated on 40 children and teenagers participants, comparing both COVID-VIRO^®^ first generation and the new COVID-VIRO ALL IN^®^. All of them found COVID-VIRO ALL IN^®^ much easier to use and much more comfortable. For young children, the COVID-VIRO ALL IN^®^ self-test appears to be safer (less risk of trauma compare to nasal swabs and no liquid exposure) easier to use than classic COVID self-tests and giving immediate result which is not the case for RT-PCR done on saliva samples (currently done in routine for kids in French schools). It could be an adapted tool for future mass screening campaigns in schools or at home under adult supervision for kids from the age of 3.

## 1 INTRODUCTION

COVID-19 is an infectious respiratory disease caused by the SARS-CoV-2 virus. As of now, this disease which started in Wuhan, China in 2019, has spread worldwide and has been acknowledged as a pandemic disease on 11 March 2020 by the World Health Organization (WHO).^1^ COVID-19 is characterised by pneumonia and upper/lower respiratory tract infection. Symptoms appear after an average incubation period of 5.2 days, the most common being fever, cough and fatigue, but also headache, sore throat and even acute respiratory distress syndrome leading to respiratory failure.^2^

Numerous highly sensitive and specific tests have been developed in an intent to control the spread of the epidemic. Nowadays, detection of SARS-CoV-2 infection is mainly executed through the RT-PCR (Reverse transcription Polymerase chain reaction) method based on the molecular detection of the virus genetic material from a nasopharyngeal sample Highly sensitive and reliable, this method still requires very specific and expensive material / equipment, as well as trained staff to perform the test in a safe and reliable manner. Results can also take several days before acquisition. Finally, although nasopharyngeal sampling is generally safe, this procedure is not exempt of any physical nor psychological risk, especially if performed in a repetitive and intensive manner.^3^ Cerebrospinal leak and meningitis have indeed been observed in some cases.^4–6^

RT-PCR saliva sampling has recently come to light as a nasopharyngeal sampling alternative especially for young children massive repeated screening campaigns in schools. It is a simpler sampling method with an estimated sensitivity of (85 % IC 95 % [82 %-88 %]. ^7,8^

Alternatively, lateral-flow immunoassays have been developed to detect the presence of the virus antigens from nasopharyngeal samples. These SARS-CoV-2 specific antigen assays (Rapid Antigen Diagnostic Test, RADT), available in pharmacy provide reliable results and were adapted for the purpose of home usage, from simple and more comfortable sampling methods (nasal sampling), enabling globally-scaled testing. Quick results from this method allows early detection and isolation of Covid-19 cases.^9^

The FDA has already approved several antigen home tests and the French health authority (Haute autorité de Santé, HAS) has defined the minimal performance requirements for these tests with an emphasis on the necessity of conducting real life studies.^10–12^

In a previous study, we demonstrated that the COVID-VIRO^®^ (AAZ-LMB, Boulogne-Billancourt, France) antigen-based rapid detection self-test was an appropriate RADT thanks to high performance and good usability assessments results, on an adult population. To further improve the test usability and thus user’s satisfaction while keeping the best performance possible, AAZ developed a new, simpler COVID-VIRO ALL IN^®^ nasal samples test where all components of the previous COVID-VIRO^®^ test are gathered in an “all in one” device.

COVID VIRO ALL IN^®^ uses a nasal sponge sampling method integrated at the end of the device. The first objective of this study was to evaluate the diagnostic performance of COVID-VIRO ALL IN^®^ on children and adults compare to the reference method (nasopharyngeal RT-PCR). The second objective was to evaluate usability of COVID-VIRO ALL IN^®^ for both children and adults, compared with the previous COVID-VIRO^®^ test.

## 2 MATERIALS AND METHODS

The study was evaluated and approved by the French ethics committee (Comité de Protection des Personnes Nord-Ouest IV) in October 2020 and was notified to the French data protection authority. This study was conducted in accordance with the Declaration of Helsinki. This implies that all participants provided written informed consent before undergoing any study-specific procedure. Two different study settings were used, one for the performance study on adults and children and one for the usability/practicability study on adults. Usability/practicability was also assessed for children, while comparing COVID-VIRO ALL IN^®^ and COVID-VIRO^®^. Consent from parents or legal representative was collected for all non-adult participants.

The performance study was set in the two COVID units of the Centre Hospitalier Régional d’Orléans: La Madeleine Hospital and La Source Hospital. The inclusion criteria for the performance study were the following: minors (ages from 5 to 17 years old) with the agreement of the legal representative or adults volunteers (> 18 years old), harboring mild to moderate symptoms (headache, fatigue, fever, sore throat, aches and pains, loss of smell and taste etc.), for less than 7 days and not requiring immediate hospitalization. The non-inclusion criteria were: hospitalised patients, symptomatic patients with symptoms duration > 7 days, asymptomatic patients or asymptomatic contact with a known case.

Regarding the usability study, the adult volunteers who participated were patients of a medical analysis laboratory (Drouot laboratory, Paris) while children participants were recruited from our infectious diseases department (Orléans Regional Hospital). No specific inclusion/non-inclusion criteria were applied.

### 2.1 In vitro diagnostic device under investigation

COVID-VIRO ALL IN^®^ (AAZ-LMB, Boulogne-Billancourt, France) is a vertical flow test that uses highly sensitive monoclonal antibodies to detect SARS-CoV-2 core antigen in a nasal sample. The test uses monoclonal antibodies to the SARS-CoV-2 core protein attached to the test area (T) on a nitrocellulose strip. A monoclonal antibody to the SARS-CoV-2 core protein labelled with colloidal gold is used as a freeze-dried conjugate.

In the test, SARS-CoV-2 antigens in the sample interact with monoclonal anti-SARS-CoV-2 antibodies to form a coloured antibody-antigen complex. This complex migrates by capillarity across the membrane to the test line (Figure 1) where it is captured by the membrane bound monoclonal anti-SARS-CoV-2 antibodies. A coloured test line appears in the results window if SARS-CoV-2 antigens are present in the sample. The intensity of the coloured test line will vary depending on the amount of SARS-CoV-2 antigen present in the sample. If no SARS-CoV-2 antigen is present in the sample, no colour will appear on the test line. The control line is used as a procedural control and should always appear in the control area if the test procedure is performed correctly. The visual interpretation of the result can be performed after 15 minutes.

**Figure 1.**
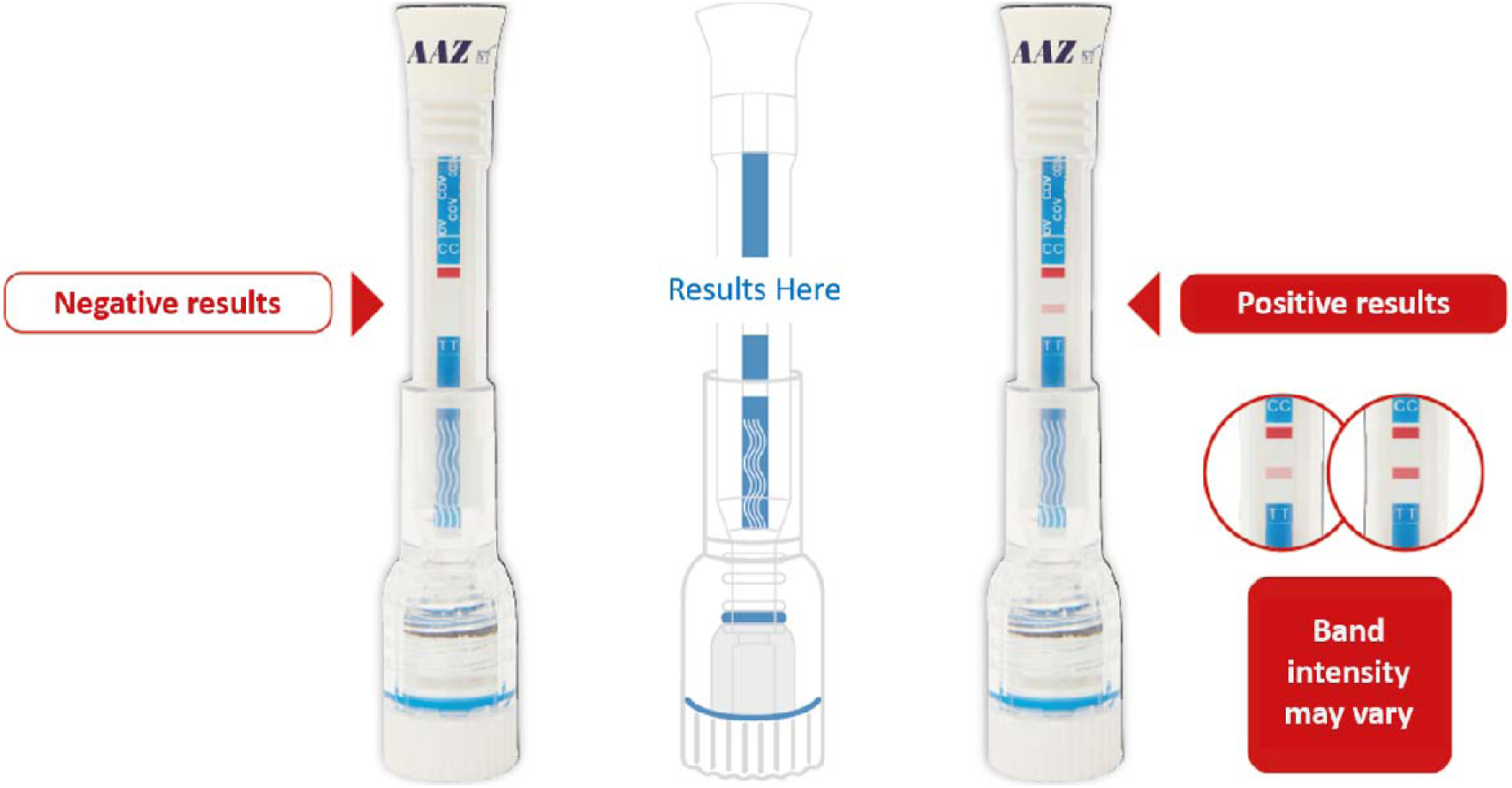
Visual appearance of the COVID-VIRO ALL IN^®^ test device and representation of the potential results

### 2.2 Comparator

The RT-PCR test for SARS-CoV-2 was performed in the virology unit of the CHR Orléans, France. Nucleic acid extraction was performed with an automated sample preparation system MGISP-960 (MGI, China). Real-time PCR detection of SARS-CoV-2 RNA targeting the ORF1ab, S and N genes was performed with the TaqPath V2 Covid-19 Multiplex RT-PCR kit (Thermofisher, Illkirch, France). Amplification was performed on QuantStudio5 (Applied Biosystems). The results of the assay were performed according to the manufacturer’s instructions. The assay includes an internal RNA extraction control and an amplification control. The samples were analysed taking into account the new positivity criteria of the French Microbiology Society’s expert committee (version 4 of 14 January 2021), in particular taking into account the specific characteristics of the Thermofisher kit used for the RT-PCR measurement.

### 2.3 Methodology

#### 2.3.1 Performance study

Upon arrival in the study centre, patients were registered for nasopharyngeal RT-PCR testing. Eligible patients were informed about the study. After consent to participate, the trained nurse performed the COVID-VIRO ALL IN^®^ nasal test and recorded the test result on the previously filled-in collection form without communicating it to the patient. Then, a nasopharyngeal swab was taken by the nurse for the RT-PCR test by the hospital laboratory. The RT-PCR test was performed using the TaqPath V2 Covid-19 Multiplex RT-PCR from Thermofisher (Thermofisher, Illkirch, France) including a variant screening. The RT-PCR result was then communicated to the patient within 24 hours and recorded in the patient’s file.

#### 2.3.2 Usability study on adults (>18 years old)

##### 2.3.2.1 Comprehension of instructions and test execution

Each participant was asked to consult the instructions for use (written, French language only, available in Figure 1) in full before carrying out the self-test. Each person was then asked to collect a nasal sample in both nostrils with the nasal sponge included in the device, dip the device back into the diluent pad, pierce the buffer capsule by pushing the device in the support, and wait for the valid result (Figure 1). Each participant was asked to comment on the different steps of the self-test on a questionnaire (Table 1). The person performing the test was supervised by an observer (laboratory staff, nurse or doctor) who gave an a posteriori assessment of the performance of the various steps by filling in an evaluation form for each participant (Table 1).

**Table 1.** COVID-VIRO ALL IN^®^ usability and supervisor’s questionnaires for Adults

| USABILITY QUESTIONNAIRE |  |  |  |
| --- | --- | --- | --- |
| QUESTION |  |  |  |
| 1. What is your opinion regarding the written instructions for the COVID-VIRO ALL IN<sup>®</sup> test? | ☐ Poor | ☐ Good | ☐ Very good |
| 2. What is your opinion regarding the execution of the nasal swab for the COVID-VIRO ALL IN<sup>®</sup> test? | ☐ Difficult | ☐ Easy | ☐ Very easy |
| 3. What is your opinion regarding the execution of the procedures COVID-VIRO ALL IN<sup>®</sup> test? | ☐ Difficult | ☐ Easy | ☐ Very easy |
| SUPERVISOR QUESTIONNAIRE |  |  |  |
| QUESTION |  |  |  |
| 1. What is your opinion on the participant's performance of this test? | ☐ Poor | ☐ Good | ☐ Very good |

##### 2.3.2.2 Interpretation of test results

The usability study also included a test result interpretation exercise during which the observer instructed the participant to randomly select 1 of 4 contrived self-tests (1 negative, 2 positive and 1 invalid), read it and give his/her interpretation of the result. The participant interpretation as well as his/her opinion on the ease of interpretation was collected on a questionnaire (Table 2).

**Table 2.** Interpretation questionnaire for Adults

| QUESTION |  |  |  |
| --- | --- | --- | --- |
| 1. What is the result of the test? | ☐ No band | ☐ 1 band | ☐ 2 bands |
| 2. How did you interpret this result? | ☐ Negative | ☐ Positive | ☐ Inconclusive |
| 3. How would you describe the reading and interpretation steps of the test? | ☐ Difficult | ☐ Easy | ☐ Very easy |
| 4. How would you describe the ease of interpretation of the COVID-VIRO ALL IN<sup>®</sup> nasal self-test? | ☐ Difficult | ☐ Easy | ☐ Very easy |

#### 2.3.3 Usability study on children and teenagers (<15 years old)

In the same fashion as the adults, each participant was asked to comment on the different steps of the COVID-VIRO ALL IN^®^ and COVID-VIRO^®^ self-tests on a questionnaire (Table 3). For 12-15 years old participants, the test was supervised by an observer (laboratory staff, nurse or doctor) who gave an a posteriori assessment of the performance of the various steps by filling in an evaluation form for each participant (Table 3). The test was directly performed by the parent/legal representative on 3-11 years old participants. The usability study also included a test result interpretation exercise during which the observer instructed the participant to randomly select 1 of 4 contrived self-tests (1 negative, 2 positive and 1 invalid, Figure 2), read it and give his/her interpretation of the result. The participant interpretation as well as his/her opinion on the ease of interpretation was collected on a questionnaire (Table 4). The participant was then asked his preferences concerning COVID-VIRO ALL IN^®^ or COVID-VIRO^®^ (Table 5).

**Table 3.**
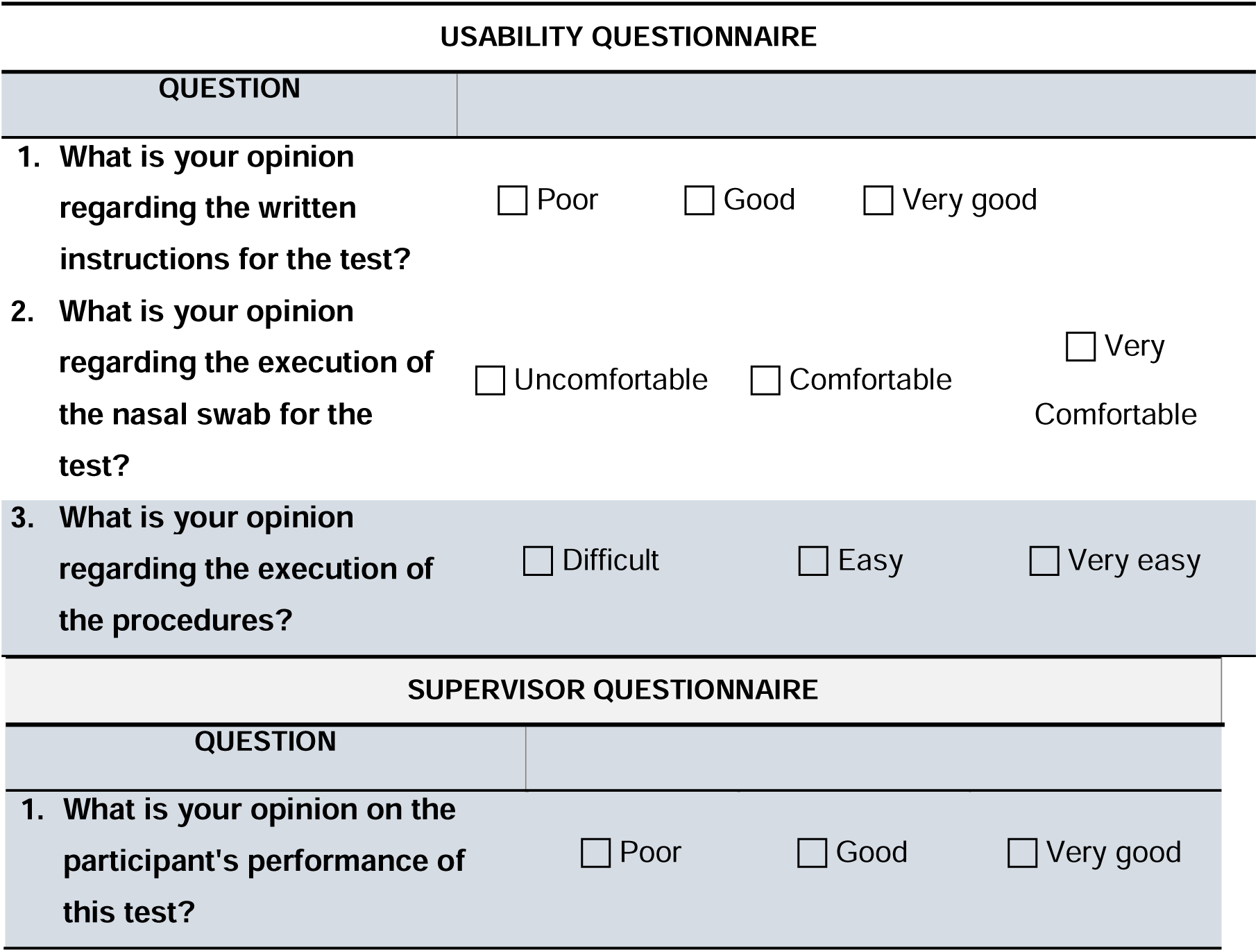
COVID-VIRO ALL IN^®^ and COVID-VIRO^®^ usability and supervisor’s questionnaires for Children/Teenagers

**Table 4.** COVID-VIRO ALL IN^®^ and COVID-VIRO^®^ interpretation questionnaire for Children and Teenagers

| QUESTION |  |  |  |
| --- | --- | --- | --- |
| 1. What is the result of the test? | ☐ No band | ☐ 1 band | ☐ 2 bands |
| 2. How did you interpret this result? | ☐ Negative | ☐ Positive | ☐ Inconclusive |
| 3. How would you describe the reading and interpretation steps of the test? | ☐ Difficult | ☐ Easy | ☐ Very easy |
| 4. How would you describe the ease of interpretation of the nasal self-test? | ☐ Difficult | ☐ Easy | ☐ Very easy |

**Table 5.** COVID-VIRO ALL IN^®^ and COVID-VIRO^®^ comparison questionnaire for Children and Teenagers

| QUESTION |  |  |
| --- | --- | --- |
| 1. Which test did you prefer? | ☐ COVID-VIRO ALL IN<sup>®</sup> | ☐ COVID-VIRO<sup>®</sup> |
| 2. Why? | Describe here |  |

### 2.4 Data analysis

#### 2.4.1 Performance study

Populations were described in terms of percentage, mean, standard deviation, range and median values. The test data was analysed in the Department of Infectious. True positive (TP) results were defined as positive individuals according to the reference method considered positive by the COVID-VIRO ALL IN^®^ test, FP (false positive) results were negative individuals according to the reference method considered positive by COVID-VIRO ALL IN^®^ test, FN (false negative) were positive individuals according to the reference method, considered negative by the COVID-VIRO ALL IN^®^ test and VN (true negative) were defined as negative individuals according to the reference method considered as negative by the COVID-VIRO ALL IN^®^ test. The specificity (Sp), sensitivity (Se), positive predictive value (PPV; probability that subjects with a positive screening test truly have the disease), negative predictive value (NPV, probability that subjects with a negative screening test truly don’t have the disease) and overall percent agreement (percentage of correctly classified instances) of the COVID-VIRO^®^ test compared to the reference test (RT-PCR) were calculated according to the following formulas:

- **Sp** (%) = 100 × [TN / (TN + FP)]
- **Se** (%) = 100 × [TP / (TP + FN)]
- **PPV** (%) = 100 × [TP / (TP + FP)]
- **NPV** (%) = 100 × [TN / (TN + FN)]
- **OPA** (%) = 100 × [(TN + TP) / (TN + FN + TP + FP)]

Confidence intervals for sensitivity and specificity were obtained with the Wilson score method.

#### 2.4.2 Usability study

Populations were described in terms of absolute number and percentage.

## 3 RESULTS

### 3.1 COVID-VIRO ALL IN^®^ Performance study

A total of 124 patients were recruited in the city of Orléans. Out of these 124 patients, only one was excluded as his RT-PCR was considered unconclusive according to the classification criteria of the French Microbiology Society^13^. Specifically, this patient was RT-PCR positive for the N gene and ORF gene but with a Ct value of 33 and 37 respectively. According to the French guideline ^13^, this sample is positive from a laboratory stand point but is excreting very low level of SARS-CoV-2 virus (Ct>32) Indicating a negligible level of contagiousness. Therefore, according to the French guideline the lab can either consider such samples weak positive or negative. As such, this participant was removed from the analysis. From the remaining 123 patients left, 4 were identified as asymptomatic patients, and thus were also excluded from this study. Consequently, the final study population comprises 119 patients. The sex ratio of the study population was 0.78 (52 men and 67 women). The median age was 37 years (mean: 38 years, range: 80 years). Among this population, the median duration of symptoms before the sampling date was 2 days (mean: 2.55, range: 7). Two groups were constituted according to the RT-PCR test results: 39 positive and 80 negative samples. The results are presented in Table 6.

**Table 6.** Performance of the COVID VIRO ALL IN^®^ antigenic rapid test in the overall Children and Adult population.

|  |  |  |
| --- | --- | --- |
|  |  | **Total** |
|  |  | **N=119** |
| **True Positive** | N | 119 |
|  | VP | **40/119 (33.6%)** |
|  |  | **Sensitivity: 93,02 %** |
| **False Positive** | N | 119 |
|  | FP | **0 (0.0%)** |
| **False Negative** | N | 119 |
|  | FN | **3/119 (2.5%)** |
| **True Negative** | N | 119 |
|  | VN | **75/119 (63.9%)** |
|  |  | **Specificity = 100 %** |
| Overall agreement | N | **119** |
|  | TP + TN | **116/119 (97,5%)** |
|  |  | **97,5 % concordance** |

In the overall population, the COVID-VIRO ALL IN^®^ test performed as follows:

- Sensitivity: 93.02% (95 %CI: 81.4% - 97.6%)
- Specificity: 100% (95 %CI: 95.2% - 100%)
- Positive predictive value: 100%
- Negative predictive value: 96.2%
- Overall percent agreement: 97,5 %

Overall agreement of results between RT-PCR and COVID-VIRO ALL IN^®^ were observed for 116 (97.5%) patients.

### 3.2 COVID-VIRO ALL IN^®^ Usability study

#### 3.2.1 Substudy 1 – COVID-VIRO ALL IN^®^: Adults comprehension of instructions, test execution and interpretation of results

A total of 68 patients (41 women and 27 men), from 18 to 67 years old (mean: 34 years) and from different occupational categories, participated in this study. None of them was excluded.

Regarding the quality of the COVID-VIRO ALL IN^®^ self-test instructions, all participants but one (98.5%) found the quality of the written instructions to be either good or very good. The last participant chose to respond “bad” to that question. Regarding the ease of execution of the COVID-VIRO^®^ sample collection (nasal sampling), 100% of the participants found it easy or very easy (16.2% and 83.8% respectively). Likewise, COVID-VIRO ALL IN^®^ self-test procedures were considered easy or very easy to perform by nearly all the participants (25.0% easy and 73.5% very easy). Only one found them hard to execute. Interestingly, it appeared that it was the same patient that was not satisfied with the written instructions. During the execution of the test, every participant was supervised by a trained professional (physician, nurse or laboratory personnel) and had the opportunity of requesting his/her assistance. Then, the supervisor had to rate the execution quality of the test procedures by the participant. Only 2/68 (2.9%) of the subjects were considered as having poorly executed the test procedures whereas 66 (97.1%) were rated as good or very good (21/68 - 30.9% and 45/68 - 66.2%, respectively). The most frequent observation was that some test users were repositioning the protective cover on top of the test after sample collection, despite instructions asking to throw it away, making it impossible to carry out the next steps properly. As an interpretation exercise, the subjects were then randomly given one out of four contrived tests and requested to read and interpret its result. Among all participants, 21 sorted a negative test, 4 an invalid test and 43 a positive test. Overall, none of the 68 patients misinterpreted the test. Only one patient found the reading and interpreting steps difficult, the majority considering them to be either easy (14.7%) or very easy (83.8%).

#### 3.2.2 Substudy 2 – COVID-VIRO ALL IN^®^ and COVID-VIRO^®^ comparison on Child and Teenager populations

##### 3.2.2.1 Comprehension of instructions and test execution

A total of 24 children (from 3 to 11 years old) and 16 teenagers (from 12 to 15 years old), with parents from different occupational categories, participated in this comparison study between COVID-VIRO ALL IN^®^ and COVID-VIRO^®^ usage and interpretation. None of them was excluded. All were followed by a supervisor who had to rate the execution quality of the test procedures, and none of them had any problem with the realisation of the test except one 10 years old participant.

Regarding the quality of both self-test instructions, nearly all participants considered the quality of the written instructions to be either good or very good. In the 3 to 11 years old population, most children found the COVID-VIRO ALL IN^®^ instructions to be very good (20/24, 83.3%), a slightly lower result was observed for COVID-VIRO^®^ instructions (17/24, 70.8%). In the teenager population COVID-VIRO ALL IN^®^ instructions seem as easier to understand (13/16 very good, 81.3%) compare to COVID-VIRO^®^ instructions (11/16 very good, 68.8%). One 14 years old participant considered the COVID-VIRO^®^ instructions poorly written, but not COVID-VIRO ALL IN^®^ instructions.

Similarly, all participants but one found both the COVID-VIRO^®^ and COVID-VIRO ALL IN^®^ sample collection rather easy or very easy. Following this, 7 out of 30 total participants (23.3%) found that the COVID-VIRO^®^ nasal swab sampling was uncomfortable, whereas only 2/30 (6.7%) felt the same way for COVID-VIRO ALL IN^®^ nasal sample collection.

Altogether, both populations found the procedures for both COVID-VIRO^®^ and COVID-VIRO ALL IN^®^ easy or very easy to perform. Only 2 teenagers found the COVID-VIRO^®^ self-test difficult to execute, and another one felt the same way about the COVID-VIRO ALL IN^®^ self-test.

##### 3.2.2.2 Interpretation of results

Just like the adult population, child and teenager populations were randomly given a contrived test to interpret the results (both a COVID-VIRO^®^ and a COVID-VIRO ALL IN^®^ self-test). Overall, there was only one child participant who misunderstood the results on one test. Otherwise, no interpretation error was observed for all other participants

Lastly, children and teenagers were asked whether they preferred the COVID-VIRO^®^ or COVID-VIRO ALL IN^®^ self-test. In the child group, 19 participants out of 24 preferred the COVID-VIRO ALL IN^®^ test (79.2%). Similarly, 14 out of 16 teenagers preferred the COVID-VIRO ALL IN^®^ test (87.5%). Overall, 33 participants out of the 40 preferred the COVID-VIRO ALL IN® (82,5%)

Participants expressed that the COVID-VIRO ALL IN^®^ system was:

- Easier to read
- Simpler to use
- More comfortable/less painful.

## 4 DISCUSSION

In our previous prospective study on COVID-VIRO^®^ rapid antigenic test diagnostic performance in real life conditions on nasopharyngeal samples^14^, the performance of the test was very similar to the reference method since the specificity and sensitivity were 100% and 96.88% respectively which placed it above the requirements of the French National Authority for Health (HAS) (sensitivity ≥ 80%, specificity ≥ 99%) ^12^ and the World Health Organization (WHO).^15^ Our current study showed COVID-VIRO ALL IN^®^ present almost same performances with 93.02% sensitivity, 100% specificity and an overall agreement of 97,5% with nasopharyngeal RT-PCR.

In addition to the performance assessment, we conducted a usability study following the FDA recommendations.^16^ The participant was asked to read the test instructions and perform all the procedures while being supervised, in order to assess whether the participant was able to correctly perform the test on his/her own and interpret it accurately. This time, 68 adults of different age, education level and socio-economic background, were included in the study constituting a representative sampling of the French general population. According to them, the quality of written instructions is good enough to be easily understood, proving that the documents provided with the COVID-VIRO ALL IN^®^ test are accessible for all laypersons.

Moreover, this study showed that the COVID-VIRO ALL IN^®^ test is very practicable since all participants were able to obtain a valid and interpretable result without requesting the supervisor’s assistance.

In terms of satisfaction about COVID-VIRO ALL IN^®^ usage, all the participants declared that the sample collection or the subsequent testing procedures were easy or very easy to perform, except one. Similarly, only one participant found the reading and the interpretation steps somewhat difficult. All together, these results demonstrate that the COVID-VIRO ALL IN^®^ test is highly adapted for use by an adult layperson.

Similar usability study was conducted on a population of 40 children (24 between 3 and 11 years old and 16 teenagers between 12 to 15 years old), while comparing directly COVID-VIRO ALL IN^®^ and COVID-VIRO^®^ first generation usage satisfaction. When asked which test they preferred, most of them (83%) answered COVID-VIRO ALL IN^®^ because it is “easier”, “simpler” or “more comfortable”. As such, the new COVID-VIRO ALL IN^®^ self-test allows a high accurate diagnostic as good as first generation COVID-VIRO^®^, but with better usability and participant satisfaction.

Following our previous study, it seems that the usability of a COVID self-test is improved with COVID-VIRO ALL IN^®^ for all types of population and especially for children, by combining all components into one easy-to-use self-device, while maintaining high performance of the test. As a RADT, it is a great alternative since it lowers the risks and adverse effects of nasal classic swabs actually used with COVID self-tests. The sampling device seems indeed well adapted for use by young children under adult supervision. The current French school screening strategy for young children is based on the RT-PCR test done on saliva. This matrix is known to be less sensitive than the nasopharyngeal sample that remains the current gold standard.^8^ In our study, the performance of COVID-VIRO ALL IN® self-test were as good as RT-PCR done on saliva samples. The ALL IN ONE self-test seems to be a potential additional tool for large testing operations and especially for young children in schools or at home under adult supervision. A recent study from Colosi et al., confirmed that weekly screening would reduce the number of cases on average by 24% in elementary and 53% in middle school compared to symptom-based testing alone^17^. This result confirms the great interest of a massive repeated screening campaigns in school based on self-testing. COVID-VIRO ALL IN could replace or help to complement actual testing strategies (saliva RT-PCR tests) and give both a reliable and quick answer to a key population (children under 12 years old) which are currently not eligible to vaccination.”

## Data Availability

The data that support the findings of this study are available from the corresponding author upon reasonable request.

## ACKNOWLEDGMENTS

The authors would like to thank the technical staff of the Department of Infectious diseases for their excellent assistance. Furthermore, the authors thank Thibaut de Sablet and Alexandre Bourgeois of Clinact, France for providing medical writing support/editorial support in accordance with Good Publication Practice (GPP3) guidelines.

## CONFLICT OF INTEREST

The authors report no conflicts of interest. The authors alone are responsible for the content and the writing of the paper.

## AUTHORS CONTRIBUTIONS

Experimental strategy design: T. Prazuck, R. Serreau, and N. G. Cassuto. Experiments: A. Gravier, M. Colin, A. Thiellay, D. Pires-Roteira and S. Pallay. Data Curation, T. Prazuck. Manuscript writing: T. Prazuck, L. Hocqueloux and R. Serreau. Manuscript editing, T. Prazuck and N. G. Cassuto.

## Notes

### Competing Interest Statement

The authors have declared no competing interest.

### Funding Statement

AAZ provided the investigational tests free of charge.
No external funding was received

### Author Declarations

COMITE DE PROTECTION DES PERSONNES NORD-OUEST IV 6, rue du Professeur Laguesse CHU LILLE CS 700001 59037 LILLE CEDEX FRANCE

